# Threshold concepts in medical education: A scoping review protocol

**DOI:** 10.1101/2021.06.18.21259141

**Authors:** Helen Jones, Lucy Hammond

## Abstract

Threshold concepts are attracting increasing attention as a framework for improving medical education practice. A growing number of studies in recent years have explored the role of threshold concepts in knowledge and skill acquisition amongst medical students and physicians. However, no review has utilised a systematic approach to examine the literature in this area. The author therefore proposes to undertake a scoping review to explore and describe the current research regarding threshold concepts in medical education and identify gaps in the existing literature. Medical and education databases will be searched for studies exploring threshold concepts in undergraduate, graduate and continuing medical education contexts. The findings will be presented in the form of a descriptive numerical summary and a narrative synthesis. The review will provide a comprehensive overview of how the threshold concept framework is currently being utilised and applied, and provide recommendations for how medical educators can employ the framework in their own practice. Exploration of the research approaches being used, and identification of gaps in the literature, will help inform future research, including determining focus for future systematic reviews.

## Background to the Topic

The idea of threshold concepts first emerged in the academic sphere in 2003 in the field of economics, where Mayer & Land (2003) built on Perkins (1999) work on troublesome knowledge to define the framework. They are described as *“akin to a portal opening up a new and previously inaccessible way of thinking about something”*, thus enabling the student to start to “think” in the manner of that discipline (Meyer & Land, 2003:1).

Threshold concepts go beyond the idea of a subject’s core concepts that are required for developing understanding, and incorporate certain unique qualities: they are transformative, in that they bring about a shift in student perception of the subject matter or in their subjective experience; irreversible, as they are difficult for the student to forget or unlearn; integrative, in that they allow students to recognise how ideas and concepts are interrelated and further their overall understanding of a subject; bounded, being delineated within a specific context or discipline; and troublesome, because they appear counter-intuitive or alien to the student, and require students to undergo the sometimes uncomfortable process of redefining previously held knowledge and understandings (Land *et al*., 2005; Meyer & Land, 2003; 2005). Crossing the threshold can lead to an elaboration of the student’s discourse and an extended use of language, described as the discursive feature of threshold concepts. In addition, cumulative learning and the subsequent shift in student identity, are the key aspects of the reconstitutive feature of threshold concepts (Meyer & Land, 2005). Whilst progressing through this transformation, students pass through a phase of liminality, in which they may shift between new and old understandings, before crossing the threshold (Cousin, 2006; Meyer & Land, 2003; 2005). Without support, students are at risk of becoming stuck in this phase, limited to a superficial understanding or ‘mimicry’, or may give up all together (Land *et al*., 2005).

The threshold concept framework therefore highlights more than just difficult topics, and their nature suggests that they can be a source of significant challenges for learning. Consequently, a recognition of threshold concepts within subject disciplines can aid in teaching and curriculum design, allowing educators to focus on them as a means of progressing student development and providing a perspective for uncovering confusion (Cousin, 2006; Land *et al*., 2005).

In medical education, it is suggested that appreciation of threshold concepts could help teachers guide students through the phase of liminality to deeper understanding of conceptually difficult topics, such as reflection, and formation of professional identify (Neve *et al*., 2016). Furthermore, it has been proposed that student understanding of the underlying theory, self-identification of threshold concepts, and reflection on their own transformative journey, may be useful in developing skills necessary for a practicing clinician (Gaunt & Loffman, 2018).

Therefore, exploration of the literature around the current understanding, application and utilisation of threshold concepts in medical education could provide important insights to guide evidence-based teaching practice. Neve *et al*. (2016) noted in their paper discussing how threshold concepts theory could inform medical education, that the framework had not been studied extensively in this context. However, pilot searching reveals there has been an increase in the number of studies published about threshold concepts in medical education since then. Exploration of the literature related to threshold concepts in health sciences education has been undertaken (Barradell & Peseta, 2017), but no reviews have taken a systematic approach to look at threshold concepts in medical education specifically. A search on 14/03/2021 of BEME, PROSPERO, JBI Evidence Synthesis, Cochrane Database of Systematic Reviews, and PubMed revealed no systematic or scoping reviews on the topic of threshold concepts in medical education.

The author therefore proposes to undertake a scoping review in order to map to current evidence. Consistent with the indications for undertaking a scoping review as stated by Munn *et al*. (2018), the purpose of this review is to identify how threshold concepts are being understood and utilised in the context of medical education, how research is being conducted in this area, and what the research gaps are. The author believes that this will contribute to the discussion around the role of threshold concepts in medical education and provide medial educators will recommendations for how they can employ the framework in their own practice.

## Aim

The aim of this study is to explore and describe the current research regarding threshold concepts in medical education and identify research gaps.

## Objective

To undertake a scoping review of literature to investigate the current extent and nature of the research about threshold concepts in undergraduate, graduate, and continuing medical education.

### Review questions

- What is the current extent and nature of the research about threshold concepts in undergraduate, graduate and continuing medical education?
- How are threshold concepts being used to inform educational practice in medical education?
- What research approaches have been used to explore threshold concepts in medical education?
- What are the research gaps in the existing literature?

## Methods

The review will follow the framework presented in the JBI Manual for Evidence Synthesis (Peters *et al*., 2020) which builds on seminal work by Arksey & O’Malley (2005) and further developments by Levac *et al*. (2010).

### Study selection criteria

#### Inclusion criteria

- **P**opulation
  - Undergraduate/graduate-entry medical students
  - Postgraduate medical trainees/residents/physicians
  - Medical educators
- **C**oncept
  - Threshold concepts
  - Fulfils at least one aspect of the taxonomy developed by Barradell & Peseta (2017) in their qualitative synthesis of threshold concepts in health science education, shown below (Table 1).
- **C**ontext
  - Undergraduate, graduate or continuing medical education
- Types of study to be included:
  - All types of paper, including empirical studies, editorials, perspectives and opinion pieces, and reviews – the decision to include all types of paper has been made in order to establish an accurate picture of the extent of the literature and current thinking in this area. The research approach will be documented in the data charting process and the implications that the type of literature has on the knowledge base will be considered in the data synthesis.

**Table 1:**
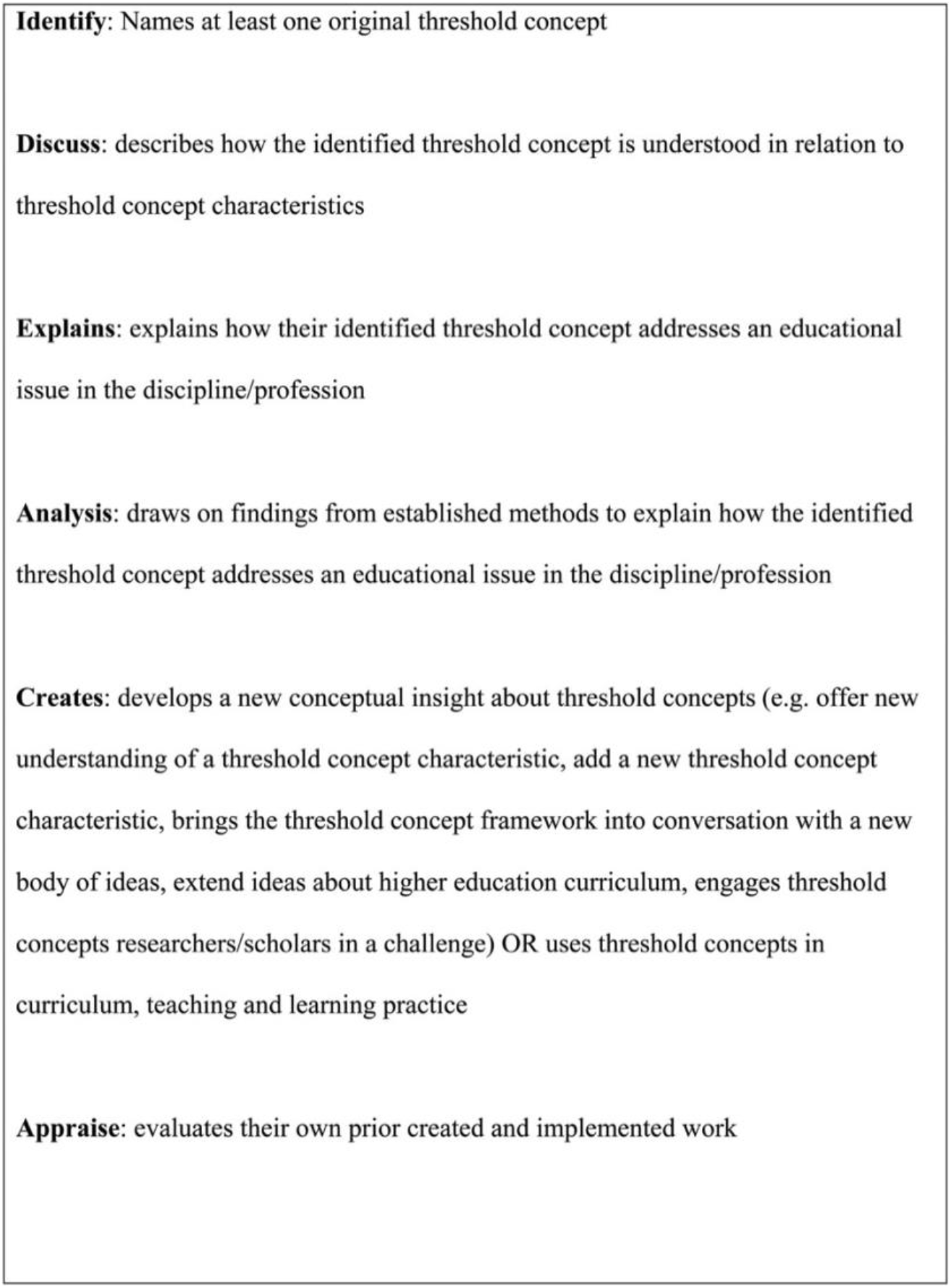
“Taxonomy of the nature of threshold concepts inquiry” from Barradell & Peseta (2017)

#### Exclusion criteria

- Studies involving other healthcare professions or health sciences, where:
  - Medical trainees/physicians/medical educators are not included
  - Or it is not possible to identify and extract data specifically related to medical trainees/physicians/medical educators
- Studies not in English language – due to time limitations and costs of translation.
- Literature which consists of an abstract only with no accompanying paper, e.g. conference abstracts – due to difficulty in charting data to a useful extent

### Search strategy

A pilot search was conducted to establish feasibility and validity, and to help identify keywords and index terms to be included in the search. The search strategy was then developed with the help of an academic librarian.

The following databases will be searched: Medline (Ovid), Embase (Ovid), Scopus (Elsevier), and Education Research Complete (EBSCO). The author will also search MedEdPublish Journals and the threshold concepts bibliography presented at https://www.ee.ucl.ac.uk/∼mflanaga/thresholds.html#thsioverv for other grey literature. The reference lists of the articles for which full-text review is undertaken will be hand searched to identify additional papers for inclusion.

The search will be limited to papers published 2003-present, to capture literature published since the first description of threshold concepts by Meyer & Land (2003). The search will also be limited to papers published in English, due to time limitations and costs of translation. All types of papers will be included, including empirical studies, editorials, perspectives and opinion pieces, and reviews. This is to ensure a broad and inclusive search, and to allow the author to establish an accurate picture of the extent of the literature and current thinking in this area. This is consistent with the approach adopted by Barradell & Peseta (2017) when exploring threshold concepts in health sciences education. However, literature which consists of an abstract only with no accompanying paper, e.g., conference abstracts, will be excluded due to difficulty in charting data to a useful extent.

The search strategy for Medline (Ovid) is shown below. The search will then be translated into the other selected databases.

Ovid MEDLINE(R) ALL <1946 to July 17, 2021>

1. exp *Education, Medical/
2. (“medical education” or “undergraduate medical education” or “graduate medical education” or “continuing medical education” or “residency” or “internship” or “clinical teaching” or “clinical education” or “medical teaching”).ti,ab,kw.
3. ((educat* or school* or university or college or curricul*) adj3 medic*).ti,ab,kw.
4. exp *Students, Medical/
5. exp *Faculty, Medical/
6. exp *Physicians/
7. ((student* or graduate* or pract* or teach* or educat*) adj3 medic*).ti,ab,kw.
8. exp *Curriculum/ or exp *Teaching/
9. 1 or 2 or 3 or 4 or 5 or 6 or 7 or 8
10. “threshold concept*”.mp. or (“threshold*” and (“transformative” or “liminal” or “troublesome” or “irreversible” or “integrative” or “bounded” or “discursive” or “reconstitutive”)).ti,ab,kw. [mp=title, abstract, original title, name of substance word, subject heading word, floating sub-heading word, keyword heading word, organism supplementary concept word, protocol supplementary concept word, rare disease supplementary concept word, unique identifier, synonyms]
11. 9 and 10
12. limit 11 to (english language and yr=“2003 -Current”)

### Screening articles

The author will follow the process outlined in Preferred Reporting Items for Systematic reviews and Meta-Analyses extension for Scoping Reviews (PRISMA-ScR) for the searching, source selection, data charting, and reporting (Tricco *et al*., 2018).

All references will be imported into Covidence (https://www.covidence.org/) and duplicates removed. The reviewers will then use Covidence to undertake the screening and review process. References will be managed in Endnote.

The two reviewers (HJ and LH) will independently screen the titles and abstracts of all articles in the search against the inclusion and exclusion criteria. If both agree the article should be included, the full text will be sought for review. If there is disagreement about whether an article should be included, the reviewers will meet to discuss and if a consensus is not reached the full text of the article will be sought and included in the next stage of the review process. In the case where articles do not contain an abstract, the full text will be sought and included in the next stage. The full texts of the selected articles will then be reviewed against the inclusion and exclusion criteria, and a final selection for the synthesis stage will be obtained. The reviewers will meet at the beginning, midpoint and end of the screening process to discuss any problems and review the search strategy if required (Levac *et al*., 2010).

### Data charting

As per the common terminology for scoping reviews, data extraction will be referred to as “data charting” (Peters *et al*., 2020). A data charting form has been developed by the author (Table 2), adapted from Peters *et al*., (2020) to capture the key details about each article and information that is relevant to the review questions. The development of the form was also informed by the data extraction table presented by Barradell & Peseta (2017) to include reference to the taxonomy utilised as part of the inclusion criteria (i.e., “Nature of inquiry”) and information that will inform the narrative synthesis (i.e., “Our findings/appraisal”).

**Table 2:** Data charting form. Adapted from Peters et al. (2020)

| **Evidence source details and characteristics** |  |
| --- | --- |
| Citation details | Author/s, date, title, journal, volume, issue, pages |
| Country in which the study conducted |  |
| Participants/ level of medical education | E.g., undergraduate medical education, postgraduate medical/resident education, continuing medical education, medical educators, mixed |
| Setting | E.g., University, hospital, community |
| Medical speciality | If stated |
| **Details/Results extracted from source of evidence** |  |
| Aims and purpose | As stated by authors |
| Research approach | E.g., type of literature (empirical study, editorial, opinion piece etc.), details of study design, data collection methods, data analysis |
| Nature of inquiry | Aspect of taxonomy and justification |
| Findings/conclusions | E.g., results of process, threshold concepts identified, outcomes of intervention, key points in discussion, conclusions drawn by authors |
| Recommendations | As stated by authors |
| Our findings/appraisal | E.g., key findings, appraisal of evidence, any limitations identified |

As recommended by Peters *et al* (2020), the data charting form was piloted by both reviewers on two articles to ensure that relevant information is charted, and changes to the form were discussed. Furthermore, data charting is an iterative process in which the data charting form is continually updated throughout the data extraction process, and this particularly the case when a review aims to explore how a theory or model has been used and applied within a study (Levac *et al*., 2010). It is anticipated that developments to the “Findings/conclusions” section of the data charting form will be made as the literature is examined and the degree of heterogenicity is established.

One reviewer will chart the data and the other reviewer will verify the data for accuracy. Any disagreements will be resolved by discussion and consensus. The extracted data will recorded as per the data charting form in Covidence. Cohen’s kappa coefficient for inter-rater reliability will not be assessed, given the iterative nature of the process and plans for resolving disagreements.

### Data synthesis

As is consistent with a scoping review, the author will not undertake a formal review of the quality or risk of bias of the selected articles (Arksey & O’Malley, 2005). However, critical appraisal of the literature will be undertaken as part of the narrative synthesis in order to address the research questions.

The data synthesis will be undertaken by the lead author. A descriptive numerical summary will be produced, including reporting of the research approaches used, the nature of inquiry into threshold concepts, and the study findings/conclusions (Arksey & O’Malley, 2005; Levac *et al*., 2010). The frequency of research approaches employed, nature of inquiry, population and contexts explored (e.g., study populations, settings, and specialities, if stated) will also be analysed and presented (Peters *et al*., 2020). The aim of this will be to map the data and identify gaps in the research. The author will then use a narrative synthesis approach to summarise and explain the main themes that emerge from the studies, correlated to the research questions (Levac *et al*., 2010). These themes may relate to the type of study or research approach, the study population or context, or the nature of inquiry into threshold concepts. However, this is an iterative process and will depend on the studies and data uncovered in the search. The aim will be to produce a descriptive account of the literature, addressing the current application of the threshold concepts framework to medical education practice, the research approaches being undertaken, and areas for future research and development.

### Project timeline

Key dates in the project timeline are shown in Table 3.

**Table 3:** Key dates in project timeline

| **Task Name** | **Start Date** | **End Date** | **Duration (days)** |
| --- | --- | --- | --- |
| Search | 17/06/2021 | 02/07/2021 | 16 |
| Screening papers (titles and abstracts) | 05/07/2021 | 30/07/2021 | 25 |
| Full-text review and data charting | 02/08/2021 | 27/08/2021 | 25 |
| Synthesis of findings | 30/08/2021 | 24/10/2021 | 55 |
| Journal article | 25/10/2021 | 09/01/2022 | 76 |

## Supporting information

PRISMA-P checklist

COI disclosure

## Data Availability

References can be found at the end of the manuscript.

## Risks and ethical considerations

No ethical review is required for this scoping review because it is limited to the review of data that is freely available in the public domain.

## Changes to the Protocol

The author does not anticipate any change to the protocol at this stage, however it is acknowledged that certain aspects of scoping reviews are iterative processes result in revisions, for example to the data charting form. Any changes will be explained and justified in the final written article.

